# Prevalence and Factors Associated with Family-Based HIV Index Case Testing in Wolaita Zone, Southern Ethiopia, 2023: A Cross-Sectional Study

**DOI:** 10.64898/2026.04.08.26350444

**Authors:** Asrat Bosha Koyra, Fatimetu Mohammed, Tewodros Eshete

## Abstract

**Background:** Family-based HIV index case testing identifies family members with unknown HIV status and links them to care. Data are limited in southern Ethiopia.

**Methods:** A facility-based cross-sectional study was conducted among 377 adults on antiretroviral therapy (ART) in Wolaita Zone, Southern Ethiopia, from November 2022 to May 2023. Participants were selected using systematic random sampling. Data were collected via interviewer-administered semi-structured questionnaire. Multivariable logistic regression identified factors associated with index case family testing. Adjusted odds ratios (AOR) with 95% confidence intervals (CI) were calculated, and statistical significance was declared at p < 0.05.

**Results:** The proportion of index case family testing for HIV was 84.9% (95% CI: 81.2– 88.6). In multivariable analysis, urban residence (AOR = 2.8; 95% CI: 1.16–6.75), duration on ART greater than 12 months (AOR = 13.0; 95% CI: 4.6–36.9), disclosure of HIV status to family members (AOR = 5.6; 95% CI: 1.9–16.5), discussion of HIV status with family members (AOR = 6.6; 95% CI: 1.9–23.2), and being counselled by health professionals to bring families for testing (AOR = 6.3; 95% CI: 2.1–19.0) were significantly associated with index case family testing.

**Conclusion:** The prevalence of family-based HIV index case testing in Wolaita Zone was 84.9%, below the national 95% target. Health professionals should strengthen counselling on ART adherence, status disclosure, family discussion, and active referral to improve testing uptake among family members of people living with HIV.

## INTRODUCTION

The human immunodeficiency virus (HIV) continues to be a major global public health concern. In 2021, an estimated 38.4 million people were living with HIV worldwide, with approximately 650,000 AIDS-related deaths and 4,000 new infections daily [1]. The Joint United Nations Programme on HIV/AIDS (UNAIDS) established the 95-95-95 targets aiming to diagnose 95% of all HIV-positive individuals, provide antiretroviral therapy (ART) to 95% of those diagnosed, and achieve viral suppression in 95% of those treated by 2030 [2]. However, an estimated 40% of people living with HIV (PLHIV) remain undiagnosed globally [3].

HIV testing and counselling is a critical gateway to ART access, which reduces morbidity, mortality, and HIV transmission through viral suppression [4]. While traditional facility-based testing approaches such as voluntary counselling and testing (VCT) and provider-initiated testing and counselling (PITC) have been widely implemented, alternative strategies are needed to reach undiagnosed populations. Index case testing—where a person with confirmed HIV infection is asked to contact family members, including children, spouses, sexual partners, siblings, and parents for HIV testing—has demonstrated higher positivity rates compared to other testing methods [5,6].

The World Health Organization recommends targeted HIV testing strategies such as index-linked testing to improve efficiency and reduce costs [7]. Index testing has proven effective in several sub-Saharan African countries, including Ethiopia, for identifying and enrolling previously undiagnosed individuals into ART [8,9]. The Ethiopian government has implemented index testing as part of its national HIV strategic plan [10].

Despite these efforts, evidence from sub-Saharan Africa, including Ethiopia, indicates that only 32–55% of adolescents are ever tested for HIV, and delayed diagnosis increases both further HIV transmission and mortality risk [11,12]. Barriers to testing include stigma, discrimination, and lack of confidential testing environments [13]. A recent population-based survey in Ethiopia found that only 40% of women had been previously tested, suggesting a substantial proportion of HIV-seropositive individuals remain unaware of their status [14].

Previous studies have identified age, residence, education, wealth, condom use, healthcare access, HIV knowledge, and substance use as factors affecting HIV testing. However, these factors vary across populations, countries, and regions [15,16]. Up to 70% of partners and family members of PLHIV are also HIV-positive, yet many remain untested [17]. Without targeted identification and testing of priority populations— including sexual partners and children of PLHIV—achieving the goal of ending HIV/AIDS by 2030 will be difficult [18].

Although index case family testing is a key strategy to identify those at highest risk, information regarding its prevalence and associated factors is limited in Ethiopia. No previous study has been conducted on family-based HIV index case testing in the Southern Nations, Nationalities, and People’s Region (SNNPR) or specifically in Wolaita Zone, one of the most populous zones in southern Ethiopia. This study addresses that gap by providing evidence-based information for policymakers, health planners, and decision-makers to improve HIV case management, reduce morbidity and mortality, and enhance service delivery.

### General objective

To assess the prevalence and factors associated with family-based HIV index case testing utilization in Wolaita Zone, SNNPR, Ethiopia, 2023.

### Specific objectives

1. To assess the prevalence of HIV index case family testing in selected healthcare facilities in Wolaita Zone.
2. To identify factors associated with family-based HIV index case testing utilization in Wolaita Zone.

## METHODS

### Study design and setting

A facility-based cross-sectional study was conducted in Wolaita Zone, Southern Nations, Nationalities, and People’s Region (SNNPR), Ethiopia. Wolaita Zone is bordered by Gamo Zone to the south, Omo River to the west, Kambata Tambaro to the northwest, Hadiya to the north, Oromia region to the northeast, Sidama region to the east, and Lake Abaya to the southeast. The administrative center is Soddo town. The zone has 63 health centers, 11 primary hospitals, 5 private hospitals, and 1 comprehensive specialized teaching hospital. The study was conducted at four selected health facilities: Wolaita Soddo University Comprehensive and Specialized Hospital, Dubbo Hospital, Soddo Health Center, and Boditi Health Center.

### Study period

Data collection was conducted from November 2022 to May 30, 2023.

### Source and study population

The source population comprised all people living with HIV (PLHIV) in Wolaita Zone who were currently receiving antiretroviral therapy (ART). The study population included all PLHIV on ART who were eligible for partner and family-based index case HIV testing services at the selected health facilities during the study period.

### Inclusion and exclusion criteria

All PLHIV on ART who were eligible for partner and family-based index case testing (i.e., those with a sexual partner or children under 15 years of age) were included. Clients who were seriously ill or physically unable to participate in an interview during the data collection period were excluded.

### Sample size and sampling procedure

The sample size was calculated using the single population proportion formula with the following assumptions: 95% confidence level (Zα/2 = 1.96), assumed proportion of eligible clients who tested their families (P = 0.49) from a previous study [19], 5% margin of error, and a total population of 3,138. The calculated sample size was 384. Since the total population was less than 10,000, a finite population correction was applied, yielding 343 participants. Adding 10% for non-response gave a final sample size of 377.

A systematic random sampling technique was used. The sampling interval (k) was calculated as 3,138/377 ≈ 8. The first participant was selected by lottery, and every 8th eligible client was subsequently enrolled. The sample was proportionally allocated across the four health facilities based on their average monthly ART patient volume.

### Data collection instruments and procedures

Data were collected using a pretested, interviewer-administered semi-structured questionnaire adapted from previous studies [19,20]. The questionnaire was developed in English, translated into Amharic, and back-translated to English to ensure consistency. It captured socio-demographic characteristics, health status, family-related factors (disclosure, discussion, partner and child testing), and health facility-related factors (counselling, trust, accessibility).

Four data collectors and two supervisors were trained for one day on study objectives, ethical principles, interviewing techniques, and data collection instruments. The principal investigator supervised data collection daily. Completed questionnaires were reviewed for completeness and consistency.

### Study variables

The dependent variable was index case family testing for HIV (yes/no), defined as either a sexual partner or biological children under 15 years having ever been tested for HIV. Independent variables included socio-demographic factors (age, sex, residence, marital status, education, occupation, income), health status factors (duration on ART, CD4 count, WHO clinical stage), family-related factors (HIV status disclosure, discussion with family members), and health facility-related factors (counselling by health workers, trust in staff, accessibility).

### Operational definitions

- **Eligible clients:** PLHIV who had a sexual partner or children under 15 years, or both.
- **Index case:** An individual newly diagnosed as HIV-positive or already enrolled in HIV treatment services [21].
- **Families tested:** Either partner or children under 15 years tested for HIV.
- **Children:** Individuals aged less than 15 years.
- **Sexual partner:** Any person with whom the index client had sexual contact at least once.

### Data analysis

Data were entered into Epi-info version 7.2 and exported to SPSS version 26.0 for analysis. Descriptive statistics (frequencies, percentages, means, standard deviations) were computed. Bivariable logistic regression was performed to identify candidate variables (p < 0.25) for inclusion in the multivariable model. Multivariable binary logistic regression using backward stepwise elimination was then conducted to identify factors independently associated with index case family testing. Adjusted odds ratios (AOR) with 95% confidence intervals (CI) were reported, and statistical significance was set at p < 0.05. Model fit was assessed using the Hosmer-Lemeshow goodness-of-fit test (p > 0.05 indicating good fit). Multicollinearity was checked using variance inflation factor (VIF < 5) and tolerance (> 0.1).

### Ethical considerations

Ethical approval was obtained from the Institutional Review Board of Saint Paul’s Hospital Millennium Medical College. Written informed consent was obtained from all participants after explaining the study purpose, procedures, risks, and benefits. Confidentiality was ensured by omitting personal identifiers. Participants were informed of their right to withdraw at any time without consequence.

## RESULTS

### Socio-demographic and health status of participants

All 377 participants completed the study (response rate 100%). Mean age was 36.6 years (SD = 9.97; range 18–75). Females comprised 52.5% (n = 198). The majority resided in urban areas (73.7%, n = 278), were married (71.1%, n = 268), and had secondary education (41.6%, n = 157). Most participants (91.2%, n = 344) had been on ART for ≥12 months (Table 1).

**Table 1.**
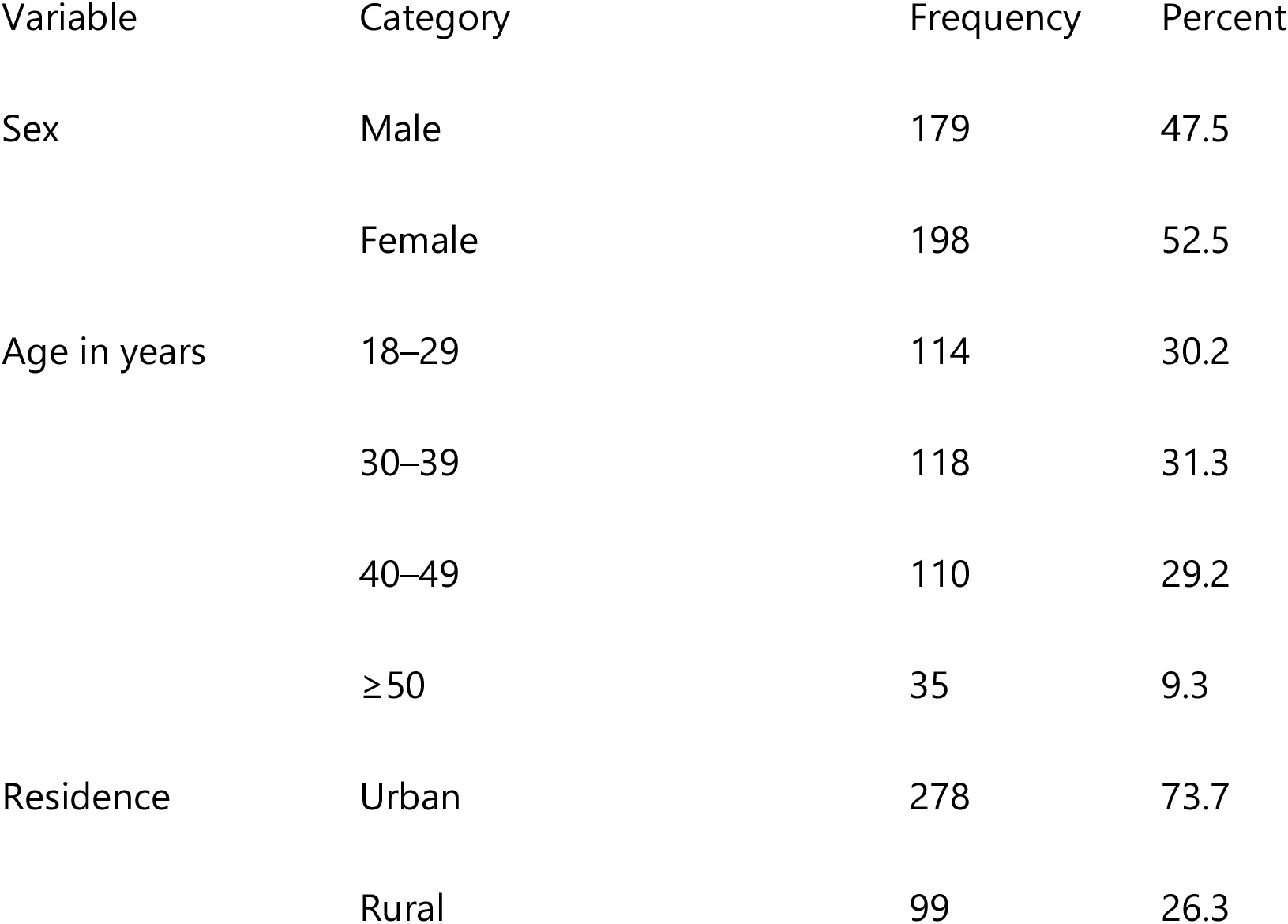
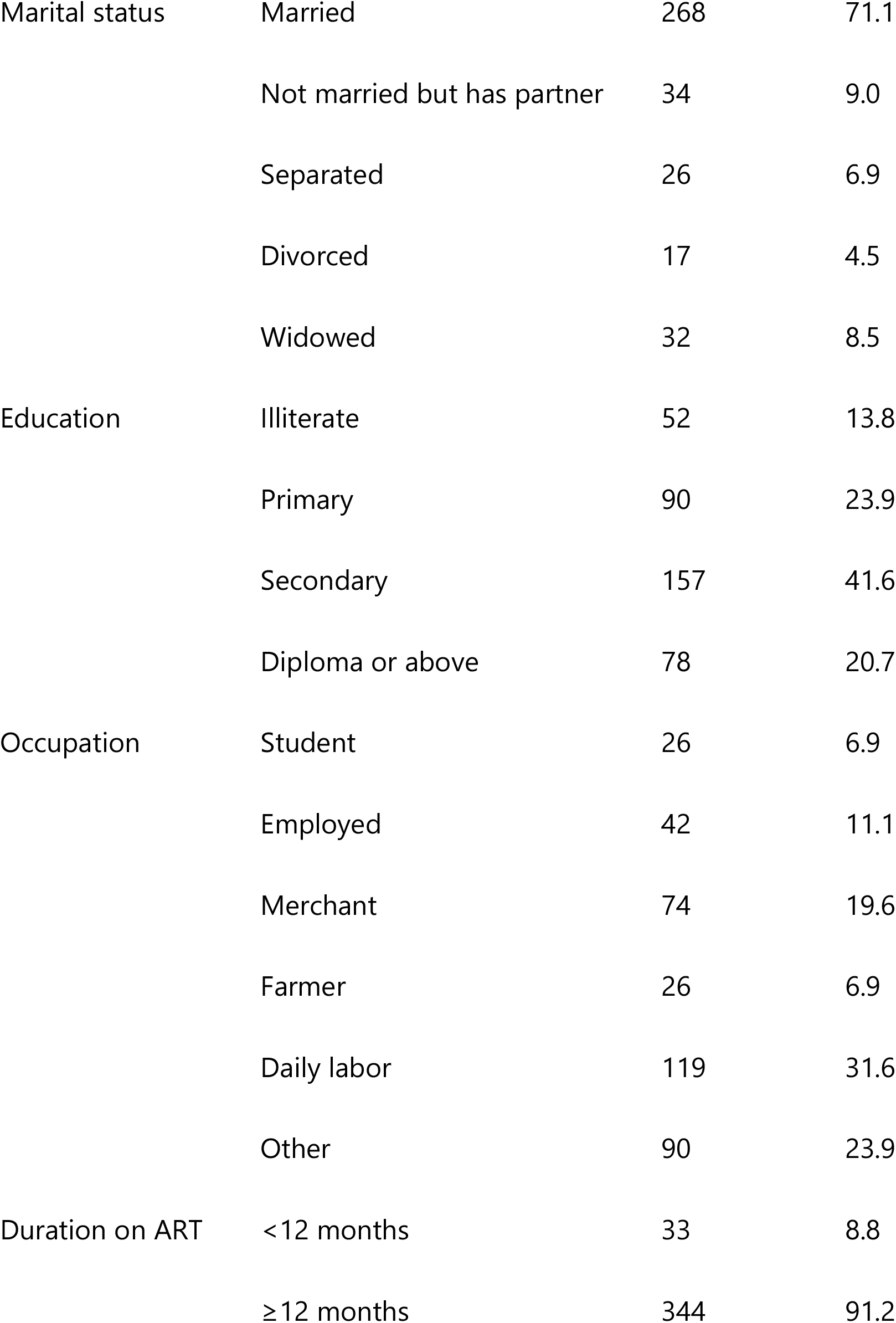
Socio-demographic and health status of index HIV cases, Wolaita Zone, SNNPR, Ethiopia, 2023 (N = 377)

| Variable | Category | Frequency | Percent |
| --- | --- | --- | --- |
| Sex | Male | 179 | 47.5 |
|  | Female | 198 | 52.5 |
| Age in years | 18–29 | 114 | 30.2 |
|  | 30–39 | 118 | 31.3 |
|  | 40–49 | 110 | 29.2 |
|  | ≥50 | 35 | 9.3 |
| Residence | Urban | 278 | 73.7 |
|  | Rural | 99 | 26.3 |
| Marital status | Married | 268 | 71.1 |
|  | Not married but has partner | 34 | 9.0 |
|  | Separated | 26 | 6.9 |
|  | Divorced | 17 | 4.5 |
|  | Widowed | 32 | 8.5 |
| Education | Illiterate | 52 | 13.8 |
|  | Primary | 90 | 23.9 |
|  | Secondary | 157 | 41.6 |
|  | Diploma or above | 78 | 20.7 |
| Occupation | Student | 26 | 6.9 |
|  | Employed | 42 | 11.1 |
|  | Merchant | 74 | 19.6 |
|  | Farmer | 26 | 6.9 |
|  | Daily labor | 119 | 31.6 |
|  | Other | 90 | 23.9 |
| Duration on ART | <12 months | 33 | 8.8 |
|  | ≥12 months | 344 | 91.2 |

### Family-related characteristics

Of 377 participants, 290 (76.9%) had disclosed their HIV status to family members, primarily to sexual partners (80.6%) and own children (51.7%). Additionally, 249 (66.0%) had discussed HIV testing with family members (Table 2).

**Table 2.**
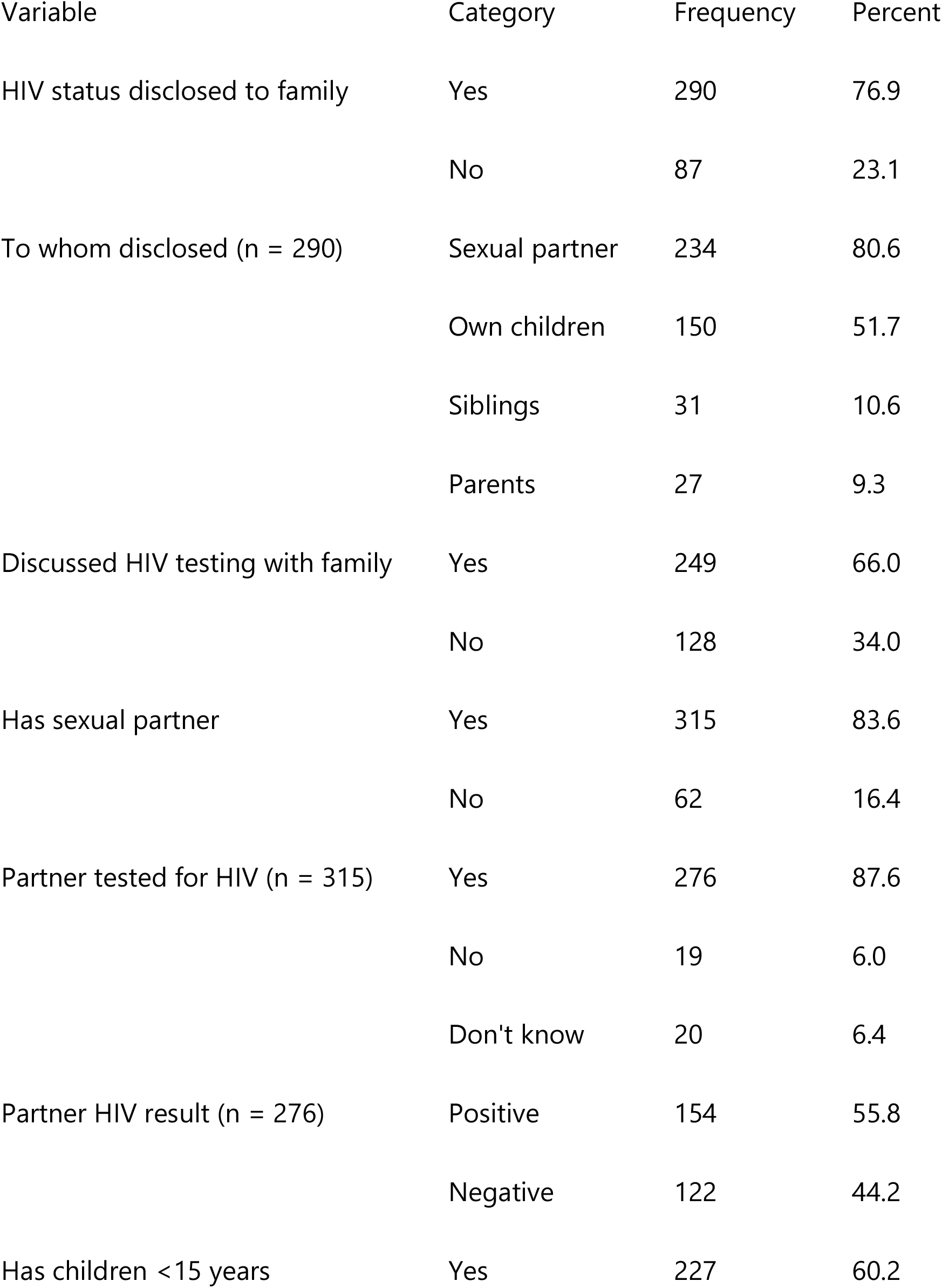
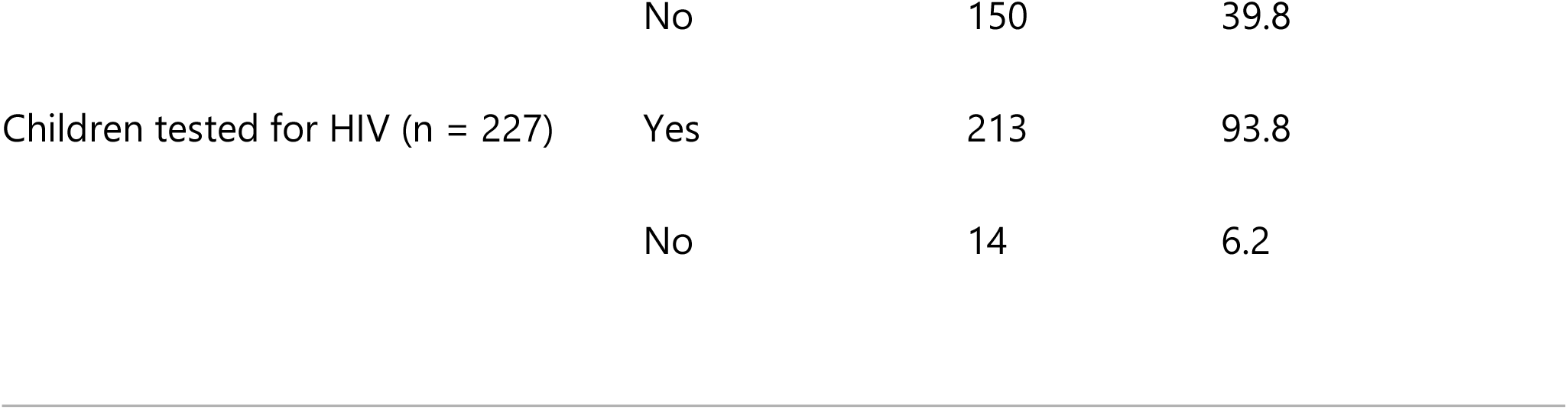
Family-related characteristics of index HIV cases, Wolaita Zone, SNNPR, Ethiopia, 2023 (N = 377)

Among 315 participants with sexual partners, 276 (87.6%) had partners who tested for HIV, of whom 154 (55.8%) were HIV-positive. Among 227 participants with children under 15 years, 213 (93.8%) had tested their children, with 48 (22.5%) having at least one child test positive (Table 2).

### Health facility-related characteristics

Most participants (96.0%) had HIV testing facilities near their residence. The majority preferred health worker referral (55.4%), health facility-based testing (93.4%), and regular work hours (87.8%). Most (92.0%) were counselled by health workers to bring families for testing, and 91.8% trusted health workers to maintain confidentiality (Table 3).

**Table 3.**
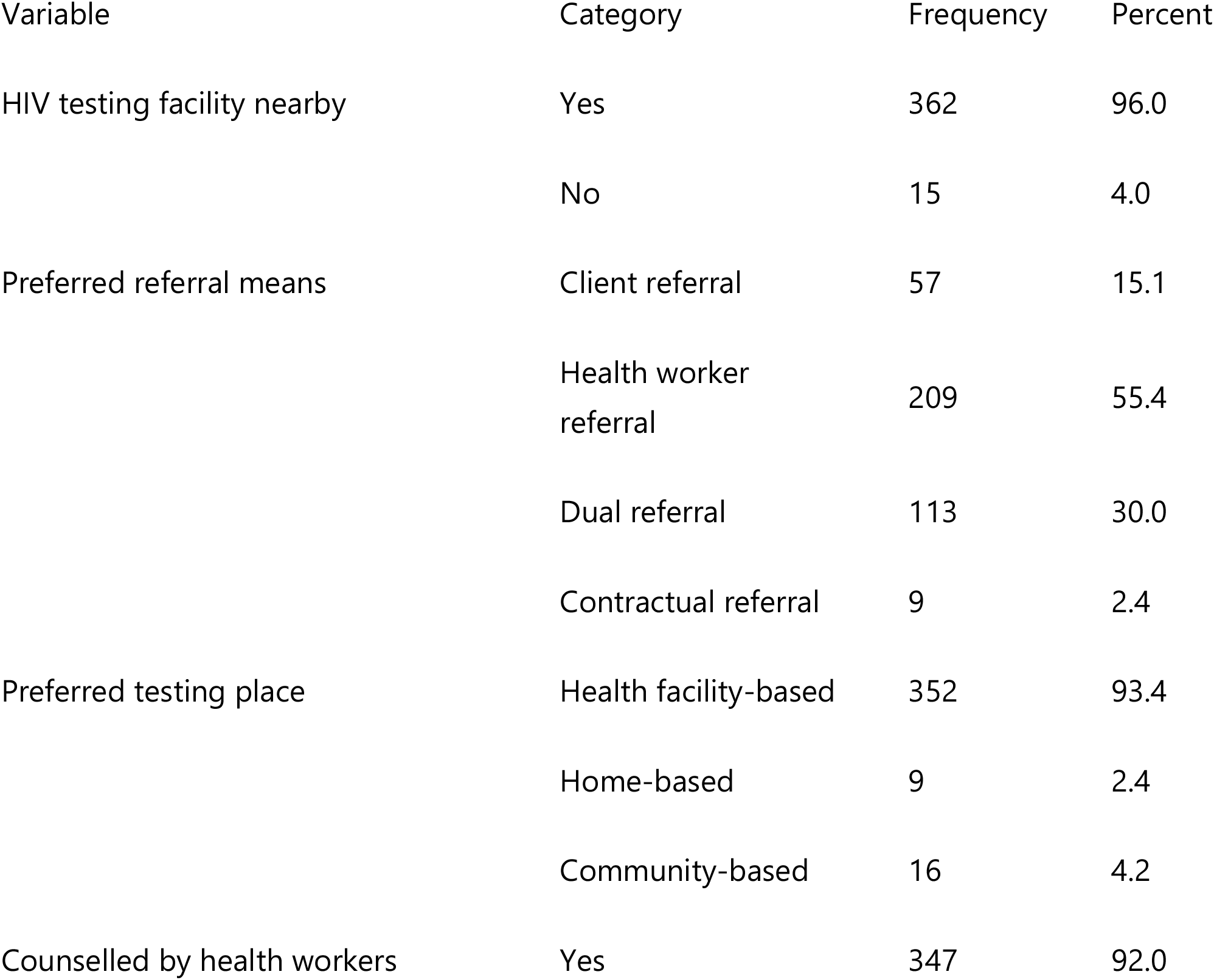
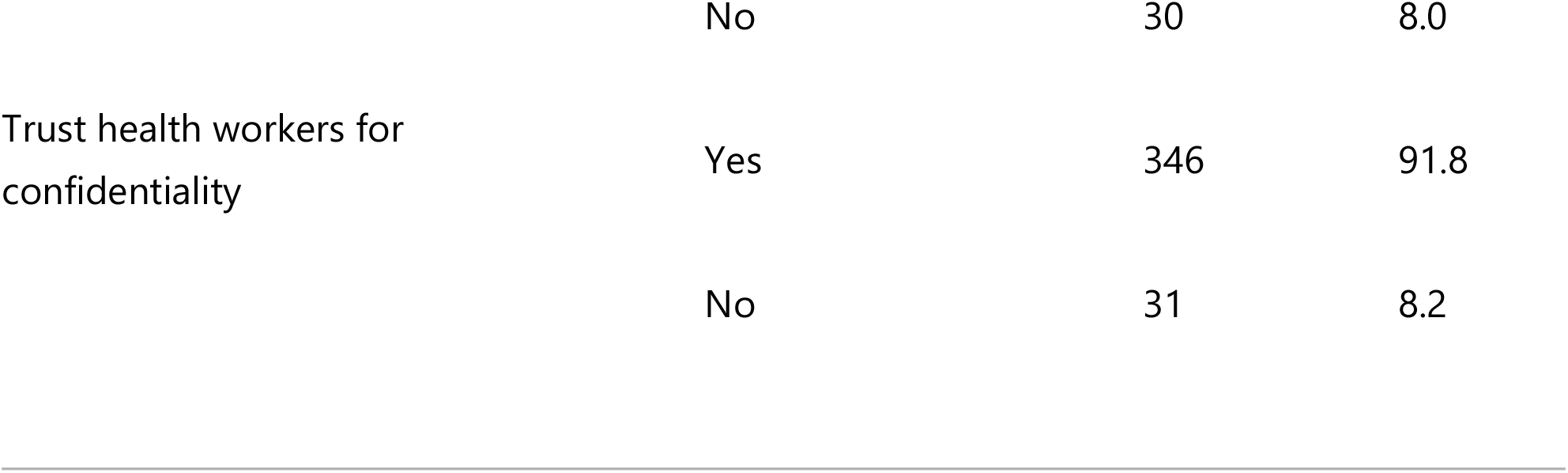
Health facility-related characteristics of index HIV cases, Wolaita Zone, SNNPR, Ethiopia, 2023 (N = 377)

### Proportion of index case family testing

Of 377 participants, 320 (84.9%; 95% CI: 81.2–88.6) had tested their family members for HIV (Figure 1).

### Factors associated with index case family testing

In bivariable analysis, residence, marital status, duration since HIV diagnosis, duration on ART, CD4 count, media exposure, HIV status disclosure, discussion with family, availability of testing facilities, trust in health workers, and counselling by health workers were candidates for multivariable analysis (p < 0.25).

In multivariable analysis, five factors remained significantly associated with index case family testing (Table 4):

**Table 4.** Bivariable and multivariable analysis of factors associated with index case family testing, Wolaita Zone, SNNPR, Ethiopia, 2023 (N = 377)

| Variable | Category | Family tested | COR<br>(95% CI) | AOR<br>(95% CI) | p-value |  |
| --- | --- | --- | --- | --- | --- | --- |
|  |  | Yes<br>(n=320) | No<br>(n=57) |  |  |  |
| Residence | Urban | 240 | 38 | 1.5<br>(0.8–2.7) | **2.8**<br>**(1.16–6.75)** | 0.023 |
|  | Rural | 80 | 19 | 1 | 1 |  |
| Duration on ART | ≥12 months | 311 | 33 | 25.0<br>(10.7–58.5) | **13.1**<br>**(4.6–36.9)** | <0.001 |
|  | <12 months | 9 | 24 | 1 | 1 |  |

| Variable | Category | Family tested | COR (95% CI) | AOR (95% CI) | p-value |  |
| --- | --- | --- | --- | --- | --- | --- |
| HIV status disclosed | Yes | 278 | 12 | 24.8 (12.1–50.7) | **5.6 (1.9–16.5)** | 0.002 |
|  | No | 42 | 45 | 1 | 1 |  |
| Discussed HIV status with family | Yes | 243 | 6 | 26.8 (11.0–64.9) | **6.6 (1.8–23.2)** | 0.004 |
|  | No | 77 | 51 | 1 | 1 |  |
| Counselled by health workers | Yes | 304 | 43 | 6.1 (2.8–13.5) | **6.3 (2.1–19.0)** | 0.001 |
|  | No | 16 | 14 | 1 | 1 |  |

\*Abbreviations: AOR, adjusted odds ratio; COR, crude odds ratio; CI, confidence interval. Bold values indicate statistical significance at $p < 0.05$ . Model fit: Hosmer-Lemeshow $\chi^2 = 3.15$ , $df = 8$ , $p = 0.67$ .\*

1. **Urban residence:** AOR = 2.8 (95% CI: 1.16–6.75)
2. **ART duration ≥12 months:** AOR = 13.0 (95% CI: 4.6–36.9)
3. **HIV status disclosure:** AOR = 5.6 (95% CI: 1.9–16.5)
4. **Discussion with family:** AOR = 6.6 (95% CI: 1.9–23.2)
5. **Counselling by health workers:** AOR = 6.3 (95% CI: 2.1–19.0)

The Hosmer-Lemeshow test indicated good model fit (χ^2^ = 3.15, df = 8, p = 0.67). No multicollinearity was detected (VIF < 5 for all variables).

## DISCUSSION

This study assessed the prevalence and factors associated with family-based HIV index case testing among adults on ART in Wolaita Zone, Southern Ethiopia. The proportion of index case family testing was 84.9% (95% CI: 81.2–88.6). Urban residence, duration on ART greater than 12 months, disclosure of HIV status to family members, discussion of HIV status with family members, and being counselled by health professionals to bring families for testing were significantly associated with index case family testing.

### Prevalence of index case family testing

The proportion of index case family testing (84.9%) is higher than reports from Kule Refugee Camp, Southwest Ethiopia (49%) [19], Felege Hiwot Referral Hospital, Bahir Dar (74.2%) [22], Uganda (72%) [23], and Kenya (62%) [24]. However, it is lower than findings from Woliso Town, Oromia, Ethiopia (94%) [20] and Tanzania (96.0%) [25]. This variation may be explained by differences in sampling methods, study populations, healthcare settings, and participant characteristics [19]. Notably, the 84.9% prevalence falls short of Ethiopia’s national 95% target for index case testing modalities [26], indicating room for improvement.

### Urban versus rural residence

Urban residents were nearly three times more likely to have tested their family members than rural residents (AOR = 2.8; 95% CI: 1.16–6.75). This finding aligns with a study in eastern Ethiopia, where urban residence was associated with 34 times higher odds of HIV testing [27]. Similarly, a multi-country study across 29 sub-Saharan African countries found consistently higher HIV testing uptake in urban areas [28]. Possible explanations include better accessibility of VCT centers, greater availability of health facilities, higher literacy rates, and reduced stigma in urban settings.

### Disclosure of HIV status

Index clients who disclosed their HIV status to family members were 5.6 times more likely to have their families tested (AOR = 5.6; 95% CI: 1.9–16.5). This is consistent with studies from Woliso, Ethiopia (AOR = 7.2) [20], Uganda [29], and Zambia [30]. Disclosure likely facilitates open communication about HIV, reduces fear and stigma, and enables family members to make informed decisions about testing. Conversely, non-disclosure—often driven by fear of stigma, rejection, or intimate partner violence— remains a major barrier [31].

### Discussion about HIV testing with family members

Participants who discussed HIV testing with their families were 6.6 times more likely to test their families (AOR = 6.6; 95% CI: 1.9–23.2). Similar associations were reported in Mekelle Hospital, Ethiopia [32], and Nairobi, Kenya [33]. Discussion creates a supportive environment, normalizes testing, and directly prompts action.

### Duration on antiretroviral therapy

Clients on ART for more than 12 months were 13 times more likely to test their families than those on ART for less than 12 months (AOR = 13.0; 95% CI: 4.6–36.9). This finding mirrors studies from Côte d’Ivoire [34], Kenya, and Uganda [35]. Newly initiated clients often experience higher levels of self-perceived stigma, fear of discrimination, and psychological distress. Over time, as clients adjust to their diagnosis, adhere to treatment, and build trust with healthcare providers, they become more willing to disclose and refer family members for testing [36].

### Counselling by health professionals

Participants counselled by health workers to bring their families for testing were 6.3 times more likely to test their families (AOR = 6.3; 95% CI: 2.1–19.0). Similar results were reported in Tanzania [37]. Effective counselling provides comprehensive information about the importance of tracing and testing sexual partners, children, and other family members. Counsellors can help PLHIV overcome fear, choose preferred referral methods, and develop practical plans for family testing [38].

### Reasons for non-testing and non-disclosure

Among participants who did not test their families, the major reasons for non-disclosure to partners included fear of perceived self-stigma (75.3%), fear of divorce or separation (24.6%), and fear of intimate partner violence (11.1%). Among those who did not disclose to children, the main reasons were not wanting to worry their children (49.3%) and believing children would not understand (32.4%). These findings highlight the need for targeted counselling interventions.

### Limitations

This study has several limitations. First, the cross-sectional design precludes causal inference. Second, data on family member testing were based on participant self-report, which may introduce social desirability and recall bias. Third, the study was conducted in only four health facilities, limiting generalizability to other settings. Fourth, HIV testing status of family members was not independently verified. Despite these limitations, this is the first study to examine family-based index case testing in Wolaita Zone and provides important baseline data for the region.

## CONCLUSIONS

The proportion of family-based HIV index case testing in Wolaita Zone was 84.9%, below the national 95% target. Urban residence, longer ART duration (≥12 months), HIV status disclosure to family members, discussion of HIV status with family members, and counselling by health professionals were significantly associated with testing uptake. Fear of self-stigma, divorce, and intimate partner violence were major barriers to disclosure and testing.

### Recommendations

- Health facilities should strengthen counselling programs that address fear and stigma, particularly for newly initiated ART clients.
- Health workers should actively counsel index cases to disclose their status, discuss testing with family members, and refer families for testing.
- Policymakers should prioritize index case testing services in all governmental and private health facilities, with special attention to rural areas.
- Further qualitative and quantitative studies are needed across different zones and regions to identify additional contextual factors.

## Data Availability

The datasets generated and/or analyzed during the current study are available from the corresponding author (Asrat Bosha Koyra) upon reasonable request.

## LIST OF ABBREVIATIONS

Abbreviation Meaning

AOR: Adjusted odds ratio
ART: Antiretroviral therapy
CI: Confidence interval
HIV: Human immunodeficiency virus
PLHIV: People living with HIV
SNNPR: Southern Nations, Nationalities, and People’s Region
SPSS: Statistical Package for the Social Sciences
VCT: Voluntary counselling and testing
WHO: World Health Organization

## DECLARATIONS

### Ethics approval and consent to participate

Ethical approval was obtained from the Institutional Review Board of Saint Paul’s Hospital Millennium Medical College. Written informed consent was obtained from all participants.

### Consent for publication

Not applicable (no individual person’s data).

### Availability of data and materials

The datasets used and analyzed during this study are available from the corresponding author upon reasonable request.

### Authors’ contributions

ABK conceived the study, designed the methodology, collected data, analyzed data, and drafted the manuscript. FM and TE supervised the study, reviewed the analysis, and critically revised the manuscript. All authors read and approved the final manuscript.

## Acknowledgements

The authors thank Saint Paul’s Hospital Millennium Medical College, the Southern Public Health Institute, Wolaita Zone Health Department, and the selected health facilities for their cooperation. We are also grateful to the data collectors, supervisors, and all study participants.

## SUPPLEMENTARY FILE

**Supplementary File 1:** Survey questionnaire (English translation). Available separately.

